# The JAK inhibitor, Tofacitinib, Corrects the Overexpression of CEACAM6 and Limits Susceptibility to AIEC Caused by Reduced Activity of the IBD Associated Gene, *PTPN2*

**DOI:** 10.1101/2024.09.26.24314341

**Authors:** Pritha Chatterjee, Vinicius Canale, Stephanie J. King, Ali Shawki, Hillmin Lei, Michael Haddad, Casey M. Gries, Dermot P.B. McGovern, James Borneman, Declan F. McCole

## Abstract

**Background and Aims:** A cohort of patients with inflammatory bowel disease (IBD) exhibit expansion of the gut pathobiont, adherent-invasive *E. coli* (AIEC). Loss of activity of the IBD susceptibility gene, protein tyrosine phosphatase type 2 (*PTPN2*), results in dysbiosis of the gut microbiota both in human subjects and mice. Further, constitutive *Ptpn2* knock-out (*Ptpn2*-KO) mice display expansion of AIEC compared to wildtype littermates. CEACAM6, a host cell surface glycoprotein, is exploited by AIEC to attach to and enter intestinal epithelial cells (IECs). Here, we investigate the role of IEC-specific *PTPN2* in restricting AIEC invasion.

**Methods:** Biopsies from IBD patients heterozygous (CT) or homozygous (CC) for the *PTPN2* SNP (single nucleotide polymorphism) *rs1893217* were processed for immunohistochemistry. HT-29 intestinal epithelial cells (IEC) were transfected with control shRNA (*PTPN2*-CTL), or a shRNA targeted towards *PTPN2* (*PTPN2*-KD). The *rs1893217* SNP was inserted (*PTPN2*-KI), or a complete knock-out of *PTPN2* (*PTPN2*- KO) was generated, with CRISPR-Cas9 gene editing of Caco-2BBe IEC lines. Adherence and invasion assays were performed with either the human IBD AIEC isolate, LF82, or a novel fluorescent-tagged mouse adherent-invasive *E. coli* (*m*AIEC^red^) at multiplicity of infection (MOI) of 10. IL-6 and the pan-JAK inhibitor tofacitinib were administered to interrogate JAK-STAT signaling. Protein expression was determined by western blotting and densitometry.

**Results:** CEACAM6 expression was elevated (colon and ileum) in IBD patients carrying the *PTPN2 rs1893217* SNP (CT, CC) compared to wildtype (TT) IBD patients. HT-29 and Caco-2BBe cell lines deficient in *PTPN2* expressed significantly higher levels of CEACAM6. Further, *PTPN2*-KI and *PTPN2*-KO cell lines also displayed greater adherence and invasion by AIEC LF82 and higher *m*AIEC^red^ invasion. CEACAM6 expression was further elevated after administration of IL-6 in *PTPN2*–deficient cell lines compared to untreated controls. Silencing of STAT1 and 3 partially reduced CEACAM6 protein expression. Tofacitinib significantly reduced the elevated CEACAM6 protein expression and the higher AIEC adherence and invasion in *PTPN2*-KI and PTPN2-KO cell lines compared to DMSO controls.

**Conclusion:** Our findings highlight a crucial role for *PTPN2* in restricting pathobiont entry into host cells. Our study also describes a role for the FDA-approved drug, tofacitinib (Xeljanz) in correcting the JAK-STAT-mediated over-expression of CEACAM6, used by pathobionts as an entry portal into host cells. These findings suggest a role for JAK-inhibitors in mitigating AIEC colonization in IBD-susceptible hosts.

## Introduction

Inflammatory bowel disease (IBD) encompassing Crohn’s disease (CD), and ulcerative colitis (UC) is a chronic, relapsing condition characterized by inflammation of the gastrointestinal tract [1–3]. IBD is a multi- factorial condition that can be caused by a faulty immune-system, environmental factors, irregularities in the host microbiome and genetic predispositions [4, 5]. One or more of these factors act in tandem to give rise to this autoinflammatory condition that features an abnormal response of the immune system to the normal intestinal flora, leading to abdominal pain, diarrhea, ulcers, and lesions. [6, 7].

Several genome-wide association (GWAS) studies have identified over 240 genes that are associated with IBD [8]. The single nucleotide polymorphism (SNP) in *rs1893217* present in the non-coding region of the gene protein tyrosine phosphatase type 2 (*PTPN2*), has been associated with several chronic inflammatory conditions like type 1 diabetes (T1D), rheumatoid arthritis (RA) and both subtypes of IBD [9, 10]. The SNP causes the loss of function in the activity of T-cell protein tyrosine phosphatase (TCPTP), the protein product of *PTPN2.* The *PTPN2* SNP *rs1893217* is carried by 19-20% of IBD patients and 16% of the general population, suggesting increased association in patient cohorts [11]. Among its substrates, PTPN2 is a negative regulator of several members of the Janus Activated Kinase – signal transducers and activators of transcription (JAK-STAT) pathway and is activated by several mediators including the pro-inflammatory cytokines IFN-γ and IL-6 [12, 13]. Several JAK inhibitors have been approved for clinical use in rheumatoid arthritis and IBD [14–16]. Tofacitinib (Xeljanz, CP-690,550) is a pan-JAK inhibitor that has been approved for use in patients with moderate to severe UC [17–19].

Along with genetic susceptibilities, altered microbial composition – dysbiosis – has been observed in many IBD patients [20]. Dysbiosis in IBD is associated with reduced microbial diversity accompanied by a decrease in beneficial commensals like Firmicutes, and a concurrent increase in Proteobacteria and Bacteroidetes [21]. *PTPN2* SNPs in IBD patients have been identified as modifiers of microbial population dynamics, as well as microbial dysbiosis and increased disease severity [33, 34]. A subset of patients also showed increased abundance of a particular B2 phylogroup of *Escherichia coli* (*E. coli*) — adherent-invasive *E. coli* (AIEC) [22]. The AIEC LF82, initially isolated from the ileal mucosa of a CD patient, had a unique ability to attach to and invade intestinal epithelial cells (IECs) and survive within macrophages [23]. Multiple groups have identified several different AIEC strains associated with both CD and UC patients, highlighting its potential role in disease pathogenesis [24, 25]. Although the exact mechanism by which AIEC contributes to IBD is still unknown, an increasing number of studies have identified its role in the maintenance and/or induction of intestinal inflammation in genetically susceptible hosts [26–29].

The IECs lining the intestinal mucosa act as a partition between the luminal bacteria and lamina propria immune cells [29, 30]. IECs also maintain a tightly regulated and selectively permeable epithelial barrier. The intestinal epithelium is also responsible for absorption of nutrients and secretion of antimicrobial peptides by enterocytes or specialized ileal crypt IECs (Paneth cells) that prevent pathogenic bacteria from penetrating the mucosal surface and provoke immune cell-mediated inflammatory responses. However, pathobionts like AIEC can utilize certain host proteins in a susceptible host to colonize the intestinal mucosa. A well-described interaction between AIEC and the human host is mediated by carcinoembryonic antigen-related cell-adhesion molecule 6 (CEACAM6) and AIEC-LF82 fimbriae protein FimH [31]. CEACAM6 is a highly mannosylated glycosyl phosphatidylinositol (GPI)-anchored protein that is expressed on the apical surface of IECs and directly interacts with AIEC to facilitate entry into epithelial cells [32].

Previously, our lab has demonstrated that *Ptpn2-*KO mice exhibit microbial dysbiosis with a profound decrease in Firmicute levels and increased abundance of Proteobacteria compared to wildtype littermate controls [33]. This increase in Proteobacteria most prominently featured a dramatic expansion of a novel mouse adherent-invasive *E. coli* (*m*AIEC) that genetically overlapped with the human AIEC LF82 [33]. Having identified that loss of PTPN2 caused an expansion of AIEC, the aim of this study was to determine the role of intestinal epithelial *PTPN2* in limiting AIEC invasion of host cells.

### Materials and methods

#### Cells

Caco-2 BBe1 cells and HT-29cl.9A IECs were cultured in Dulbecco’s modified Eagle’s Medium (DMEM; Corning, Tewksbury, MA) and McCoy’s 5A medium (Corning, Tewksbury, MA), respectively. The medium was supplemented with 10% heat-inactivated fetal bovine serum (FBS; Gibco, Waltham, MA), 1% L-glutamine (Invitrogen, Carlsbad, CA), and 1% penicillin/streptomycin (Corning, Tewksbury, MA) at 37°C incubator maintained at 5% CO_2_/air mix. Cells were cultured in 6-well plates for protein assay, or in 12-well plates for immunofluorescence. Insertion of SNP *rs1893217* (*PTPN2*-KI) and complete knockout of *PTPN2* (*PTPN2*-KO) in Caco-2 BBe cell lines was performed using CRISPR-Cas9 gene editing by Synthego (Menlo Park, CA). For *PTPN2* knockdown in HT-29 cells, lenti-viral particles containing scrambled shRNA (*PTPN2*CTL) or PTPN2-specific shRNA (*PTPN2*-KD) were generated as previously described [34].

For STAT1 and STAT3 silencing, the cells were transfected with previously validated, STAT1 and STAT3- specific, or non-targeting control siRNA constructs (Dharmacon, Lafayette, CO) using DharmaFECT transfection agents. In experiments with STAT1 and STAT3 siRNA challenged with IL-6, the culture media was replaced with serum-free medium 8 hr prior to addition of IL-6 (50 ng/ml; Peprotech, Cranbury, NJ). 24 hours post IL-6 addition, cells were washed with PBS and collected with RIPA as previously described.

In experiments with tofacitinib, the cells were treated with tofacitinib (50 μM, MedChemExpress, Monmouth Junction, NJ). Control cells were treated with an equal amount of vehicle (dimethyl sulfoxide, DMSO, 0.5%, Sigma-Aldrich).

#### Immunofluorescence

Caco-2BBe IECs were seeded at 200,000 on coverslips in 12-well plates. After 24 hours, medium was deprived of serum and antibiotics. For bacterial fluorescence assay, cells were washed with PBS (x3) followed by fixation with 4% paraformaldehyde (PFA) for 20 min at room temperature. Cells were then permeabilized with 0.3% Triton X-100 (Fischer Scientific, Waltham, MA) for 5 min followed by blocking with 5% Bovine Serum Albumin (BSA; Fischer Scientific, Waltham, MA) for 10 min at room temperature. Cells were incubated with Alexa Fluor 488-Phalloidin antibody (1:1000) (Abcam #176553, Cambridge, MA) for 90 min at room temperature followed by nuclei staining with 4,6-diamididino-2-phenylindole (DAPI; Vector Laboratories, Newark, CA). For CEACAM6 staining, cells were fixed with 4% PFA and permeabilized with Triton X-100 for 30 minutes, followed by blocking with 5% BSA for 1 hour. Cells were then incubated anti- CEACAM6 antibody (1:100; Abcam #78029; Cambridge, MA) at 37°C for 3 hours. Cells were washed with 0.1% tween supplemented PBS (X3) after which they were incubated in Alexa-Fluor 488 (1:100; #711-586- 154Jackson Research, West Grove, PA) for 1 hour. Images were captured using Leica DM5500 microscope attached with a DFC365 FX camera using a 63X oil immersion objective, or with an inverted Zeiss Airyscan. DAPI was visualized using a 405 nm excitation laser and DAPI filter set. mCherry was visualized using a 561 nm excitation laser and mCherry filter set. Images were analyzed using ImageJ software (NIH).

#### Protein isolation and Western blotting

For protein isolation from cells, the cells were washed with ice cold PBS and lysed in radioimmunoprecipitation assay (RIPA) buffer containing phosphatase and protease inhibitors (Roche, South San Francisco, CA). All samples were then sonicated for 30 seconds, centrifuged (10 min. at 13000 G at 4°C), and the supernatant transferred into fresh tubes. Protein concentration was detected using a BCA assay (Thermo Fisher Scientific, Waltham, MA). For Western blots, aliquots with equal amounts of protein were separated by electrophoresis on polyacrylamide gels, and the proteins blotted on nitrocellulose membranes. The membranes were then incubated in blocking buffer (5% milk, 1% BSA in tris-buffered saline with 0.5% Tween) for 1 hr. and incubated overnight at 4°C with appropriate antibody.The next day, the membranes were washed in tris-buffered saline with 0.5% Tween, incubated with HRP-coupled secondary antibodies (Jackson Immunolabs, West Grove, PA), washed again, and immunoreactive proteins detected using ELC substrate (Thermo Fisher Scientific, Waltham, MA) and X-ray films (GE Healthcare, Chicago, IL)

#### Immunohistochemistry of patient biopsies

Microscopy slides with paraffin-embedded and formalin-fixed (PEFF) intestinal biopsies from *PTPN2* genotyped patients (rs1893217) were provided by Dr. Dermot McGovern at Cedars Sinai Medical Center, Los Angeles, CA, USA. Twelve samples from colon (6 WT, 5 Het, 1 KO), and 12 samples from ileal segment (6 WT, 6 KO) were used in this analysis (Supplementary Table 1) [34]. Slides were deparaffinized, rehydrated and processed for immunohistochemistry as described before. Briefly, heat-induced antigen retrieval was performed for 20 minutes at ∼96 C with sodium-citrate (pH 6) buffer. Primary antibody (Abcam #ab78029, Cambridge, UK) diluted in PBS with 5% NDS at 1:200 dilution was incubated overnight at 4°C. Detection was done using the biotin-streptavidin detection system and signal developed by DAB reaction according to manufacturer’s protocol (Cell Signaling Technology #8059, Beverly, MA,). Then, sections were counterstained with hematoxylin and slides were mounted with Permount® and visualized on a Leica microscope model DM5500B with DFC450C camera (Leica – Nussloch, Germany).

#### Statistics

Data are represented as the mean of a series of ‘n’ biological repetitions ± standard deviation (SD). Data followed a Gaussian distribution and variation was similar between groups for conditions analyzed together. Differences between groups were analyzed with one-way or two-way ANOVA. Tukey post-hoc test determined *P-values. P*-values below 0.05 were considered significant. No data points were excluded from statistical analysis. Statistical analysis was performed using GraphPad Prism version 9 (GraphPad, San Diego, CA).

## RESULTS

### Loss of PTPN2 increases CEACAM6 expression in intestinal epithelial cells

We previously demonstrated that constitutive *Ptpn2-*KO mice displayed microbial dysbiosis and the expansion of adherent-invasive *E. coli* (AIEC) [33]. To understand how loss of *PTPN2* in intestinal epithelial cells affects susceptibility to AIEC, we conducted an RNA sequencing analysis comparing control (*PTPN2*- CTL) and *PTPN2*-KD HT-29cl.9A IEC cell lines (Figure 1A). The AIEC-binding receptor, *CEACAM6*, was the most significantly increased gene in *PTPN2*-KD cells compared to control cells (Figure 1A, B). We further confirmed that CEACAM6 protein levels were also significantly elevated in *PTPN2*-KD HT-29 cells compared to the control cells (Figure 1C). To demonstrate the translational significance of our *in vitro* findings, we next determined the CEACAM6 expression in the intestinal biopsies of IBD patients genotyped for *PTPN2* loss-of-function SNP *rs1893217*. IBD patients with “TT” wildtype allele were considered controls while, patients with heterozygous “CT” and homozygous “CC”, where “C” is the minor allele carrying IBD associated SNP, displayed elevated CEACAM6 expression in the colon of the IEC membranes (Figure 1D; Supplementary Table 1). Together, these data suggest that loss of functional epithelial *PTPN2* causes intestinal epithelial over-expression of the AIEC receptor protein, CEACAM6, both *in vitro* and in genotyped IBD patients.

**Figure 1.**
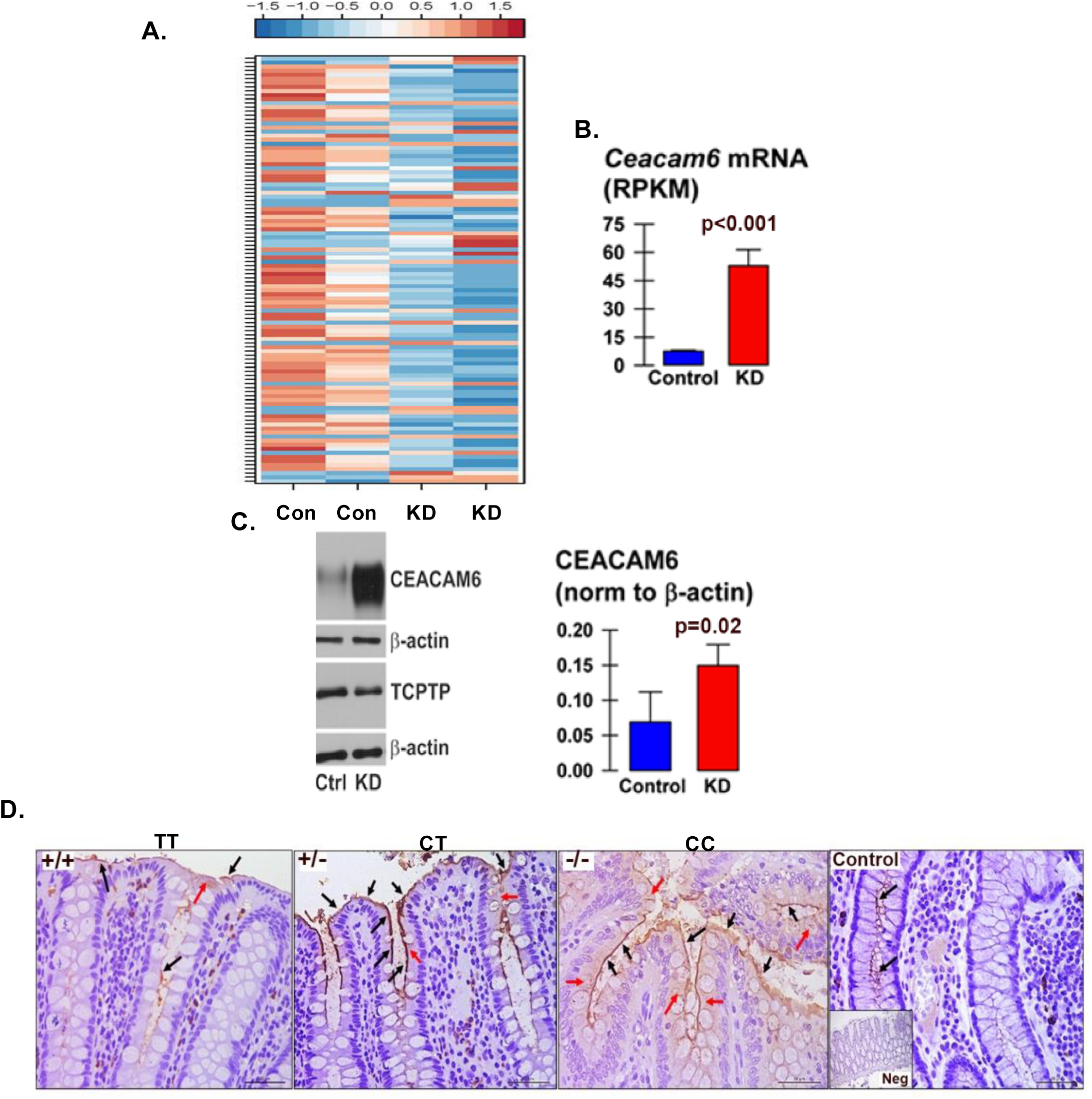
Loss of Epithelial *PTPN2* promotes CEACAM6 protein expression. **A.** RNA-sequencing analysis from *PTPN2*-CTL and *PTPN2*-KD HT-29 cells shows higher *Ceacam6* mRNA in the KD condition compared to the control condition. **B.** Quantification of the *Ceacam6* mRNA. **C.** Western blots from control and *PTPN2*-KD HT-29 cells confirm higher CEACAM6 protein expression in the KD cells. **D.** Colonic biopsies from IBD patients homozygous for the wildtype allele (TT), heterozygous (CT) or homozygous (CC) where ”C” allele possess the *PTPN2* SNP rs1893217 that is associated with IBD susceptibility. The patient biopsies from variant carrying allele showed higher expression of CEACAM6 (brown).

### Loss of Epithelial *PTPN2* Increases Susceptibility to AIEC Invasion

Next, we assessed the functional consequences of increased CEACAM6 in *PTPN2*-deficient cells. To do so, we first generated Caco-2BBe1 cell lines carrying the IBD patient associated SNP *rs1893217* (*PTPN2*- KI) cells and a complete knock-out of the *PTPN2* gene (*PTPN2*-KO) by CRISPR-Cas9 gene editing. As observed in HT-29 cells, we confirmed that expression of CEACAM6 was significantly higher in *PTPN2* deficient (PTPN2-KI and *PTPN2*-KO) IECs compared to control Caco-2 BBe cells (*PTPN2*-WT) (Figure 2 A, B). Interestingly, expression of CEACAM6 in *PTPN2*-KI cell lines was also higher than the *PTPN2*-KO cells (Figure 2 A, B). Further, we also stained for CEACAM6 expression in these cells to determine its localization within cells. We confirmed the overexpression of CEACAM6 observed in *PTPN2-*KI and KO cell lines was primarily on the cell surface albeit some of its expression was also intracellular in all 3 genotypes (Figure 2 C). Next, we determined if the increase in CEACAM6 in the KI and KO cells increased susceptibility to AIEC infection. We observed that both *PTPN2-*KI and KO cells displayed increased adherence and invasion by the human AIEC, LF82, compared to the *PTPN2*-WT cells (Figure 2 D, E). Next, we visually confirmed higher invasion of *PTPN2*-KI and KO cells by using a constitutive mCherry fluorescent tagged *m*AIEC^red^ (Figure 2 F). Here we observed some intestinal epithelial cells in all three genotypes were highly invaded by *m*AIEC^red^ as compared to other cells. This phenomenon was more apparent in *PTPN2*- KI and KO cells compared to the control cells. We therefore quantified cells that demonstrated less than 50 bacteria per cell as having “low-level” invasion, while cells that displayed more than 50 bacteria/cell were designated as having a “high level” of invasion. *PTPN2*-KI and *PTPN2*-KO cells displayed a significant increase in susceptibility to both parameters of AIEC invasion compared to the control cell lines (Figure 2 G). We also confirmed increased AIEC LF-82 adherence and invasion in *PTPN2*-KD HT-29 IECs compared to controls (Supplementary Figure 1 A, B). The *PTPN2*-KD HT-29’s also demonstrated higher *m*AIEC^red^ invasion compared to *PTPN2*-CTL cells (Supplementary Figure 1 C). Next, we confirmed a functional role for CEACAM6 in AIEC invasion. We pre-treated cells with a CEACAM6 blocking antibody. Anti-CEACAM6 antibody significantly reduced *m*AIEC^red^ invasion of *PTPN2*-KI and KO cells compared to untreated controls (Figure 2 H). We also visually confirmed that anti-CEACAM6 reduced *m*AIEC^red^ invasion of epithelial cells (Figure 2 I). Taken together, these results demonstrate loss of *PTPN2* promotes AIEC invasion of host cells by promoting CEACAM6 protein expression.

**Figure 2.**
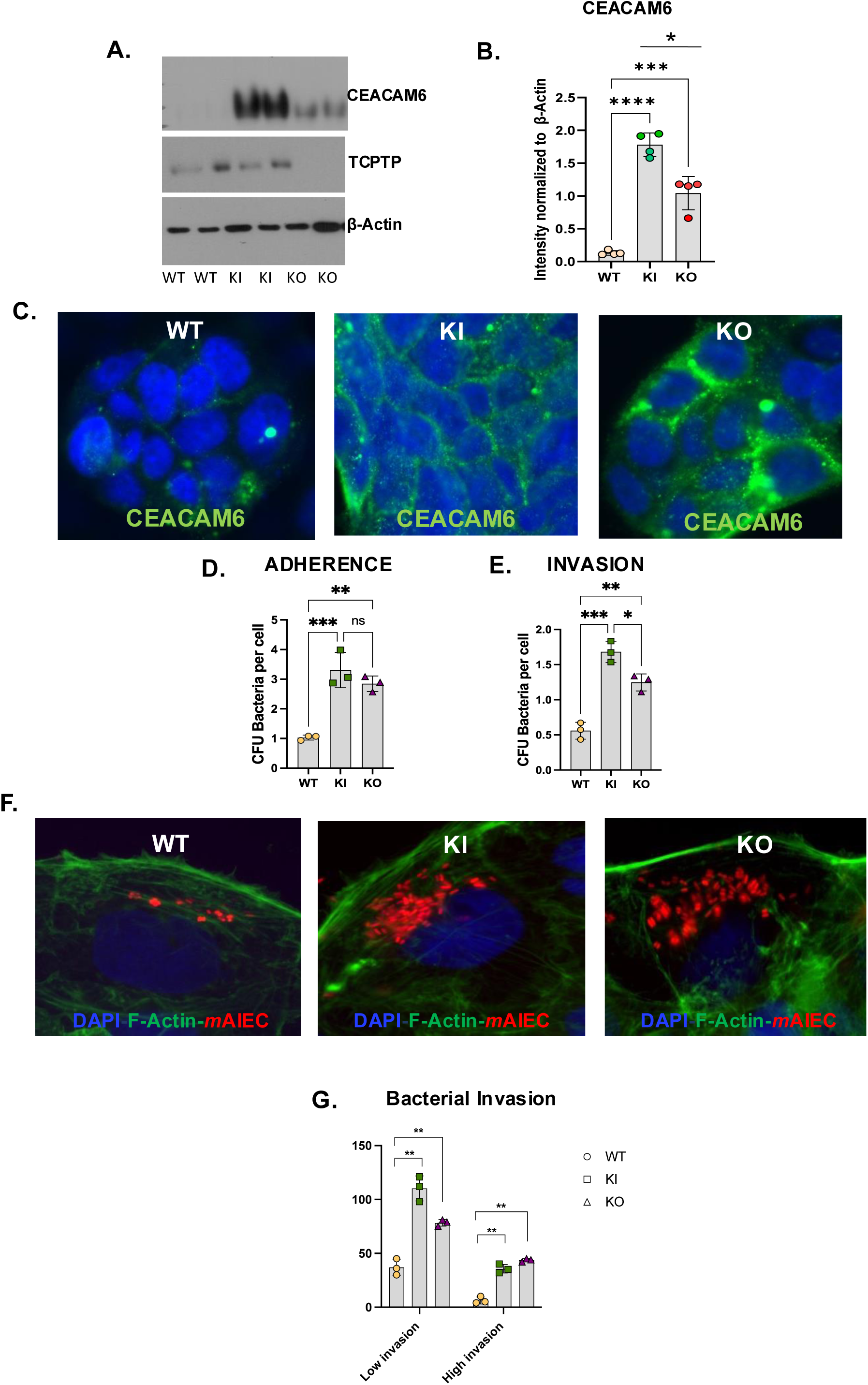
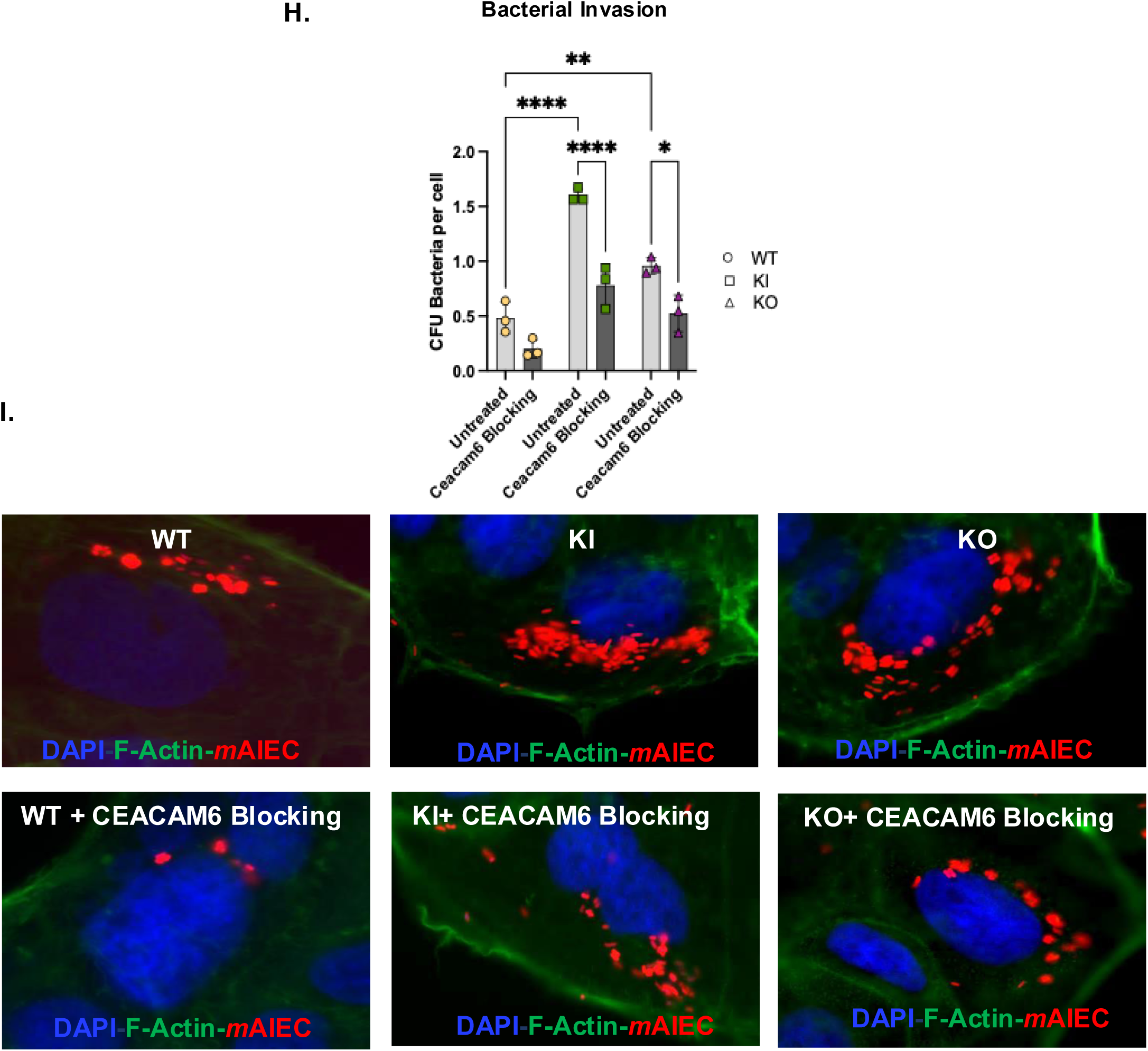
Loss of *PTPN2* in IECs increases susceptibility towards AIEC adherence and invasion. Caco-2 BBe cells were modified with CRIPSR-Cas9 to carry the *PTPN2* SNP *rs1893217 (PTPN2*-KI) or a complete knockdown of *PTPN2* gene was generated *(PTPN2*-Ko). **A**. Representative western blot showing elevated CEACAM6 expression in *PTPN2*-KI and *PTPN2*-KO cells compared to *PTPN2*-WT cells. **B.** CEACAM6 expression is significantly higher in *PTPN2*-KI and *PTPN2*-KO cells compared to WT cells. *PTPN2* KI cells have a higher CEACAM6 expression compared to *PTPN2* KO cells. *PTPN2*- WT, KI and KO cells were stained for CEACAM6. **C.** Increased apical CEACAM6 in *PTPN2* KI and KO cells is confirmed visually. AIEC LF-82 shows higher **D.** Adherence and **E.** Invasion of *PTPN2*-KI and KO cells compared to controls. **F.** Immunofluorescence showing increased *m*AIEC^red^ invasion of *PTPN2*- KI and KO cells compared to control. **G.** *PTPN2*-KI and KO cells display both high and low *m*AIEC^red^ invasion cell types compared to *PTPN2*-WT cells. CEACAM6 antibody was used to block CEACAM6 protein. After which the bacterial invasion protocol was followed as described. **H.** We observed reduced AIEC invasion in both KI and KO cells after CEACAM6 blocking **I.** Reduced AIEC invasion after CEACAM6 blocking was confirmed by immunofluorescence. *P* < 0.05 = *; *P*<0.01= **; *P* <0.005 =*** with Tukey post-hoc test.

### IL-6 Promotes CEACAM6 expression in *PTPN2*-Deficient IECs

Previous studies have shown that CEACAM6 expression can be elevated by pro-inflammatory cytokines and that it encodes a STAT3 binding site in its promoter region [37] (Supplementary Figure 4A). Since *PTPN2* is a negative regulator of the JAK-STAT signaling pathway and suppresses IL-6 induced activation of JAK-STAT signaling, we assessed whether IL-6 promotes CEACAM6 expression in epithelial cell lines. Expectedly, 24 hours post IL-6 treatment both *PTPN2*-KI and *PTPN2*-KO cells displayed higher levels of phosphorylated STAT3 compared to WT cell lines (Figure 3A). We also observed that CEACAM6 expression was elevated in *PTPN2*-WT cells after IL-6 treatment and this effect was further exacerbated in *PTPN2*-KI and *PTPN2*-KO cells (Figure 3 A, B). To determine if the elevated CEACAM6 levels were a consequence of higher STAT3 activation, we performed STAT3 silencing using siRNA constructs. Interestingly, we did not observe a reduction in CEACAM6 levels after STAT3 siRNA transfection although p-STAT3 levels were dramatically lowered in *PTPN2*-deficient cell lines (Supplementary Figure 4B). Next, we silenced both STAT1 and STAT3 in these cell lines. We observed a decrease in CEACAM6 expression between untreated and IL-6 treated *PTPN2*-KO IECs after STAT1 + STAT3 silencing (Figure 3C). We also observed a decrease in CEACAM6 expression of WT cells treated with IL-6 (control siRNA vs. STAT1 + STAT3 silencing), but no changes were observed in *PTPN2*-WT cells treated with control (scrambled) or STAT1 and STAT3 siRNA. Interestingly, no impact was observed in PTPN2-KI cells line’s expression of CEACAM6 after STAT1+STAT3 siRNA treatment (Figure 3C). We also confirmed that IFN-γ-STAT1/STAT3 induction elevated the expression of CEACAM6 (Supplementary Figure 3A, B). Together these data suggest that cytokine-induced CEACAM6 expression is regulated, at least in part, by STAT1 and STAT3 activation, and this mediates the elevated CEACAM6 expression observed in IECs with no PTPN2 activity.

**Figure 3.**
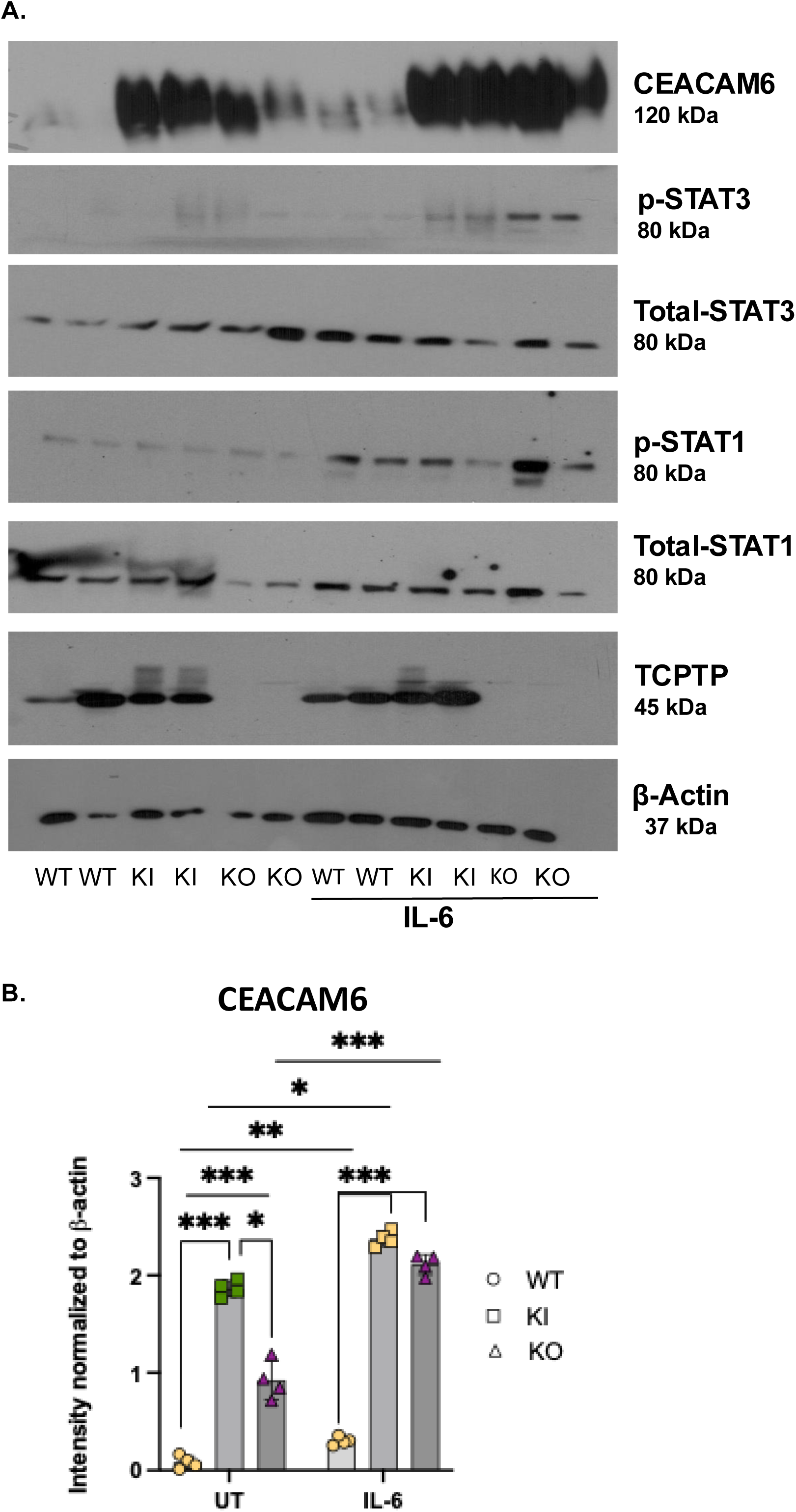
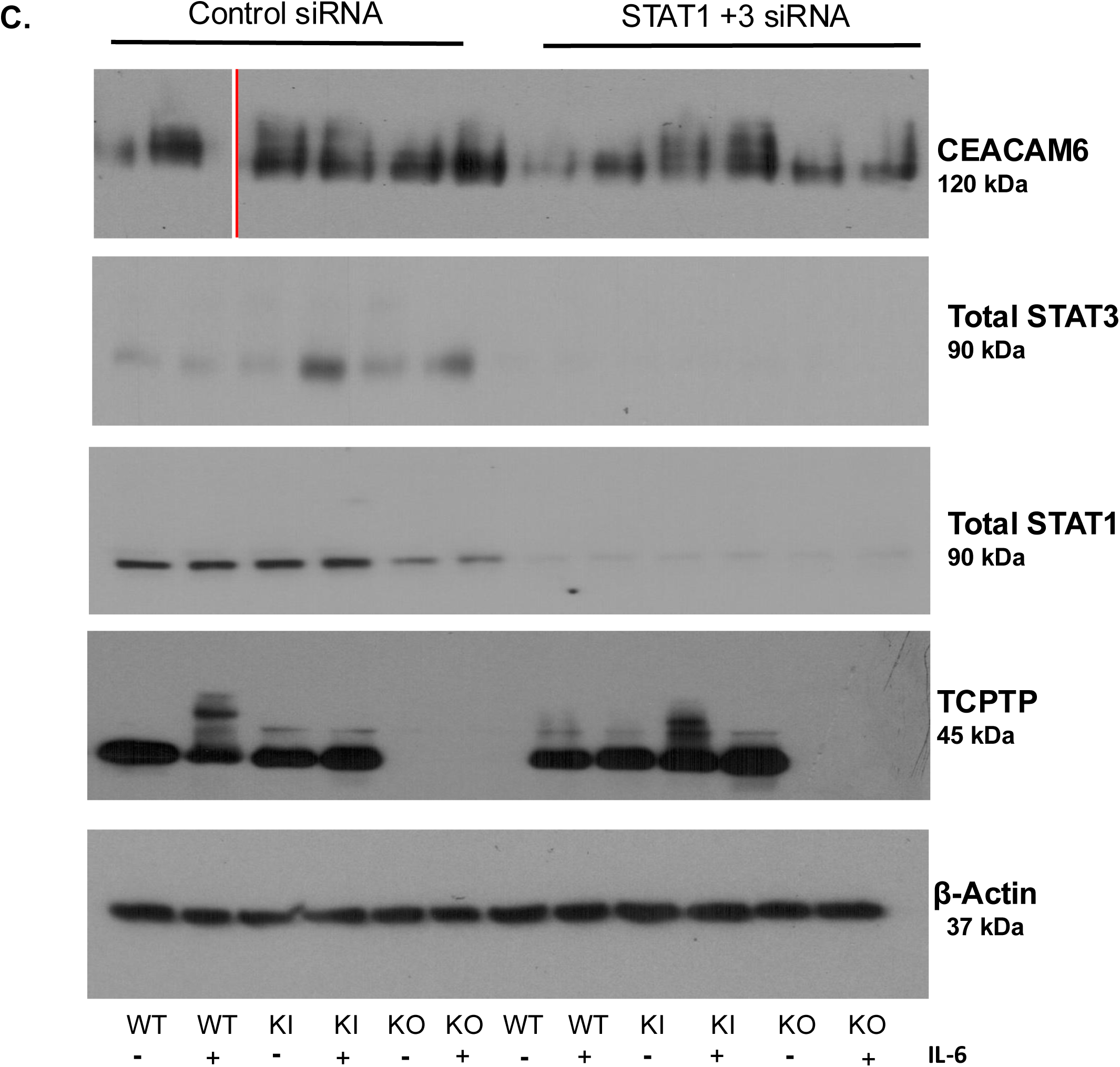
IL-6 Promotes CEACAM6 Expression in *PTPN2*-deficient Caco-2 BBe cells. *PTPN2 -* WT, KI and KO were treated with IL-6 at 50ng/ml for 24 hours. **A.** Representative western blot images depicting β-actin, PTPN2, CEACAM6, total and phosphorylated STAT1 and STAT3. CEACAM6 protein expression was elevated at basal states for *PTPN2*-KI and KO cells and was further elevated after IL-6 treatment. Phospho-STAT3 levels in *PTPN2*-KI and KO cells were also elevated after IL-6 treatment. **B.** Densitometry analysis of CEACAM6 protein expression 24-hours after administration of IL-6. CEACAM6 protein expression is significantly increased in IL-6 treated compared to untreated controls. WT, KI and KO cells were treated with non-targeting control (siControl) and STAT specific (siSTAT1) and STAT3 specific (siSTAT3) siRNA. After 72 hours, IL-6 was administered. **C.** Representative western blot images showing reduced total STAT1 and total STAT3. CEACAM6 protein expression is reduced between PTPN2-KO control siRNA and STAT1+3 siRNA. Further, CEACAM6 protein expression is reduced in *PTPN2*-WT +IL-6 treated control siRNA and STAT1 and 3 siRNA treatments.

### Tofacitinib Rescues CEACAM6 Protein Overexpression and Limits AIEC Invasion in PTPN2- Deficient IECs

Given that the inhibition of STAT 1 and 3 reduced the over-expression of CEACAM6 in *PTPN2*-KO cell line, we next determined if the FDA approved JAK inhibitor for UC, tofacitinib, could also prevent CEACAM6 over-expression and prevent susceptibility to AIEC infection. All three IEC genotypes were pre-treated with tofacitinib for 2 hours and challenged with IL-6 for another 2 hours. We observed that IL-6 increased CEACAM6 expression in all 3 genotypes (Figure 4A, B). Treatment of the KI and KO cell lines with tofacitinib drastically reduced the phospho-STAT1 and STAT3 levels both in basal and IL-6 treatment conditions (Figure 4A). Additionally, pre-treatment of *PTPN2*-KO cell lines with tofacitinib reduced CEACAM6 expression in basal and IL-6 stimulated overexpression of CEACAM6 (Figure 4 A,B). The elevated CEACAM6 in *PTPN2* KI cell lines displayed a moderate, but statistically significant, reduction in CEACAM6 protein expression with tofacitinib both in control and the IL-6 challenge condition (Figure 4A, B). Tofacitinib reversed IL-6 induction of CEACAM6 expression in PTPN*2*-WT cells compared to its respective DMSO control condition (Figure 4A, B). Interestingly, PTPN2-WT cells treated with tofacitinib, displayed increase in CEACAM6 protein expression (Figure 4A, B). We also visually confirmed that that tofacitinib reduced CEACAM6 expression in *PTPN2*-KI and KO cell lines (Figure 4C). Next, we determined if treatment with tofacitinib also reduced susceptibility to AIEC infection in *PTPN2-*deficient cell lines. We observed that *PTPN2-*WT cells showed no change in AIEC adherence or invasion in the presence of tofacitinib. However, both *PTPN2*-KI and KO cells show reduced AIEC adherence and invasion in the presence of tofacitinib (Figure 4 D, E). Together, these results suggest that tofacitinib can correct the overexpression of CEACAM6 and increased AIEC invasion found in cells with reduced PTPN2 activity.

**Figure 4.**
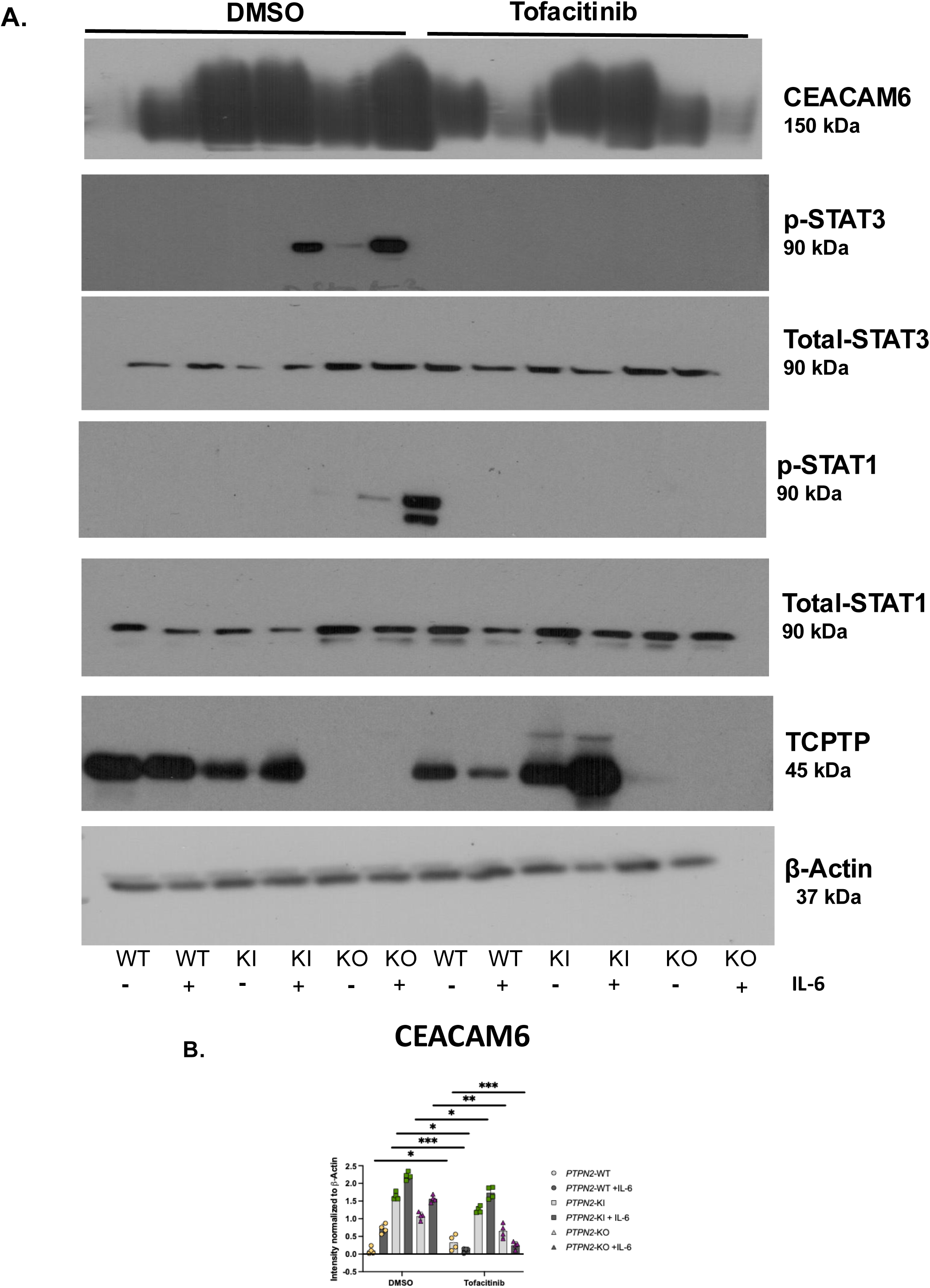
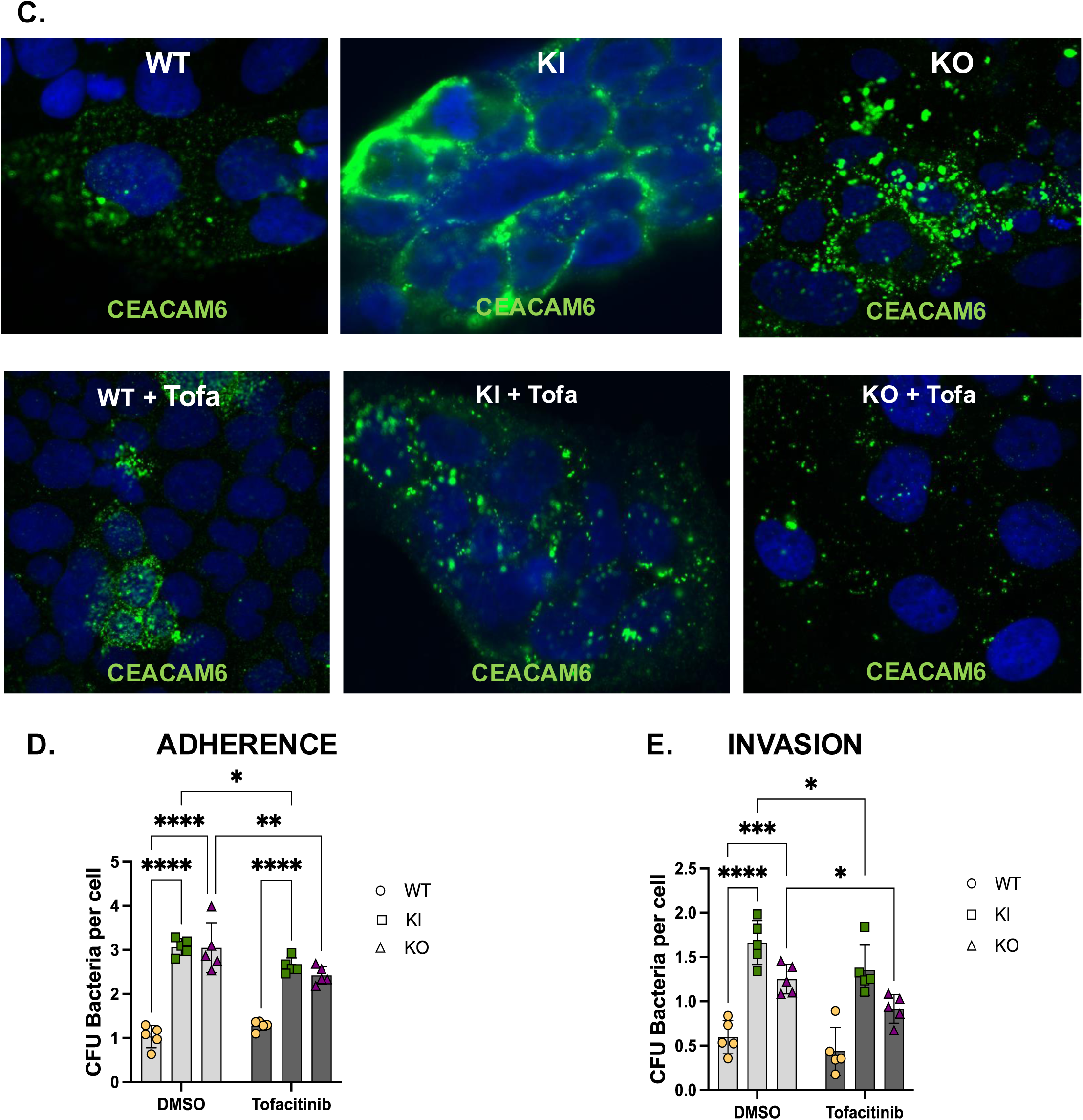
Tofacitinib Rescues CEACAM6 Over-expression and Limits AIEC Invasion in PTPN2 Deficient IECs. Cells were pre-treated with tofacitinib at 50μg/ml for 2 hour and challenged with IL-6 at 50 ng/ml for 2 hours. **A**. Representative western blot images depicting β-actin, TCPTP, CEACAM6, phosphorylated and total STAT1 and STAT3. **B.** Densitometry analysis shows CEACAM6 protein levels significantly decreased in tofacitinib pre-treated *PTPN2*-WT cells challenged with IL-6. *PTPN2*-KI cells also show moderate reduction in CEACAM6 protein levels after tofacitinib treatment. *PTPN2*-KO cells show drastic reduction in the expression of CEACAM6 with tofacitinib treatment and IL-6 challenge. **C.** CEACAM6 staining in PTPN2-WT,KI and KO cells treated with DMSO or tofacitinib treated. Reduced CEACAM6 expression after tofacitinib treatment. **D.** Adherence **E.** Invasion of AIEC-LF82 is reduced in *PTPN2*-KI and KO cell lines after tofacitinib treatment compared to DMSO controls. No effect was seen in *PTPN2* –WT cells.

## DISCUSSION

Previously, our lab has determined that loss of PTPN2 in mice causes expansion of the pathobiont, AIEC [33]. In this current study, we describe that the loss of epithelial PTPN2 increases susceptibility to AIEC invasion by increasing expression of CEACAM6, a critical receptor for AIEC that mediates entry into IECs. Further, we have proposed a unique role of the UC approved drug, tofacitinib (Xeljanz) in alleviating CEACAM6 expression in *PTPN2*-deficient cell lines and limiting susceptibility to AIEC invasion. Taken together, these data highlight a multifactorial role for *PTPN2* in maintaining microbial homeostasis and restricting bacterial entry into host cells.

In this study, we showed that CEACAM6 is significantly upregulated in *PTPN2* deficient colonic epithelial cell lines, where PTPN2 expression was stably knocked down, or CRISPR engineered to express the clinical *PTPN2* rs1893217 loss-of-function variant, or in which PTPN2 was deleted. Further, IBD patients heterozygous (CT) or homozygous (CC) for the *PTPN2* susceptibility variant *rs1893217,* displayed increased CEACAM6 expression in the ileum and colon compared to wildtype (TT) controls. A complicating feature of AIEC is that it is difficult to distinguish them from other *E. coli* pathotypes as a molecular marker unique to AIEC is yet to be identified. The field has therefore relied on phenotypic adherence, invasion assays and macrophage survival assays to functionally designate an *E. coli* as an AIEC [36–38]. Given these constraints, no large-scale study has yet identified expansion of AIEC in IBD patients carrying *PTPN2* SNPs. Moreover, it is not possible to study the *PTPN2* regulation of CEACAM6 using an *in vivo* murine model since there is no murine homologue of CEACAM6. Considering these limitations, we generated Caco-2BBe cell lines carrying the disease-relevant *PTPN2* variant *rs1893217* or in which PTPN2 was deleted. We validated that the increased expression of CEACAM6 is of functional consequence by demonstrating that *PTPN2*-variant carrying (*PTPN2*-KI), or PTPN2 deletion (*PTPN2*-KO), cell lines have increased AIEC adherence and invasion burden. Moreover, suppression of increased phosphorylated STAT1 and STAT3 partially reduced the over-expression of CEACAM6 in *PTPN2*- WT and KO cell lines challenged with IL-6, thus functionally confirming the mechanism by which reduced *PTPN2* activity increases CEACAM6 expression. Interestingly, a reduction of CEACAM6 expression was not observed in the *PTPN2*-KI cell lines, indicating that other pathways maybe involved in these cell lines that potentiate the overexpression of CEACAM6. Further translational relevance was supplied by showing that pretreatment with the JAKi, tofacitinib, reduced CEACAM6 expression in *PTPN2*-KO cells. We observed *PTPN2*-KI cells have substantial CEACAM6 expression which was marginally reduced by tofacitinib. These differences in the suppression of CEACAM6 by a JAKi, in *PTPN2*-KI vs. *PTPN2*-KO cell lines may be due to differences in the mechanisms of CEACAM6 regulation in the two genotypes. It is possible that CEACAM6 over-expression is controlled by pathways other than JAK-STAT signaling pathway in the *PTPN2-*KI cells and therefore, its expression was not drastically reduced with tofacitinib. The potential involvement of additional transcription factors governing CEACAM6 regulation in *PTPN2*-KI cells will be an area of interest for our future studies.

In murine models, our lab has demonstrated that mice lacking PTPN*2* in macrophages (*Ptpn2*-LysMCre) are more susceptible to AIEC invasion and exhibit increased bacterial translocation to extra-intestinal organs [40]. Further, CEACAM6 blocking antibody prevented AIEC invasion of human *PTPN2*-deficient macrophages demonstrating that CEACAM6 also played a role in AIEC uptake in macrophages as well. Further we have also demonstrated that mice with IEC-specific knockout of *Ptpn2* (*Ptpn2*^ΔIEC^) display higher AIEC tissue burden and increased intestinal barrier permeability compared to control groups (Supplementary Figure 2A, B -only for reviewers) (To be submitted in a separate manuscript). Altogether, our findings have identified an important role for PTPN2 in limiting AIEC pathophysiology.

CEACAM6 can be upregulated by several inflammatory cytokines and AIEC itself can upregulate CEACAM6 expression to promote its own tissue colonization [31]. Despite its high association with disease, very few therapeutic strategies have been investigated to prevent AIEC colonization. Recently, a study demonstrated that the anti-TNF medication, adalimumab, restricted AIEC replication in CD macrophages by inducing Flotillin (FLOT-1) and restricting chitinase 3-like 1 proteins (CHI3L1), an AIEC receptor, in macrophages [41]. In this study, for the very first time, we have demonstrated how an IBD-susceptibility PTPN2 gene variant regulates cell surface proteins to mediate AIEC colonization. We further demonstrate that the consequences of reduced PTPN2 activity can be mitigated with the FDA approved JAK inhibitor, tofacitinib. These data shed further light on additional cellular and mechanistic targets of JAKi that may contribute to overall therapeutic benefit. Our findings further suggest that tofacitinib, and potentially other JAK1 or JAK3 targeting drugs may prove particularly effective in treating patients carrying loss-of-function *PTPN2* SNPs not only with respect to promoting mucosal barrier healing, but also to restrict pathobiont colonization.

**Table 1:** Colonic tissues from CD patients harboring the loss-of-function IBD-associated *PTPN2* rs1893217 SNP. Adapted from: Marchelletta, R. R., M. Krishnan, M. R. Spalinger, T. W. Placone, R. Alvarez, A. Sayoc- Becerra, V. Canale, A. Shawki, Y. S. Park and L. H. Bernts (2021). "T cell protein tyrosine phosphatase protects intestinal barrier function by restricting epithelial tight junction remodeling." The Journal of Clinical Investigation 131(17).

| Variant | Diagnosis | Region | Sex | Age Range |
| --- | --- | --- | --- | --- |
| AA/TT | CD | Colon | F | 45-50 |
| AA/TT | CD | Colon | M | 20-25 |
| AA/TT | CD | Colon | F | 40-45 |
| AA/TT | CD | Colon | F | 70-75 |
| AA/TT | CD | Colon | F | 30-35 |
| AA/TT | CD | Colon | F | 20-25 |
| GA/CT | CD | Colon | M | 70-75 |
| GA/CT | CD | Colon | F | 15-20 |
| GA/CT | CD | Colon | F | 45-50 |
| GA/CT | CD | Colon | F | 45-50 |
| GA/CT | CD | Colon | F | 70-75 |
| GG/CC | CD | Colon | M | 30-35 |
| AA/TT | CD | Ileum | F | 45-50 |
| AA/TT | CD | Ileum | M | 20-25 |
| AA/TT | CD | Ileum | F | 40-45 |
| AA/TT | CD | Ileum | F | 70-75 |
| AA/TT | CD | Ileum | F | 30-35 |
| AA/TT | CD | Ileum | F | 20-25 |
| GG/CC | CD | Ileum | F | 45-50 |
| GG/CC | CD | Ileum | M | 25-30 |
| GG/CC | CD | Ileum | F | 35-40 |
| GG/CC | CD | Ileum | M | 25-30 |
| GG/CC | CD | Ileum | F | 35-40 |
| GG/CC | CD | Ileum | M | 40-45 |

## Data Availability

All data produced in the present work are contained in the manuscript.

---

**Supplementary Figure 1:**
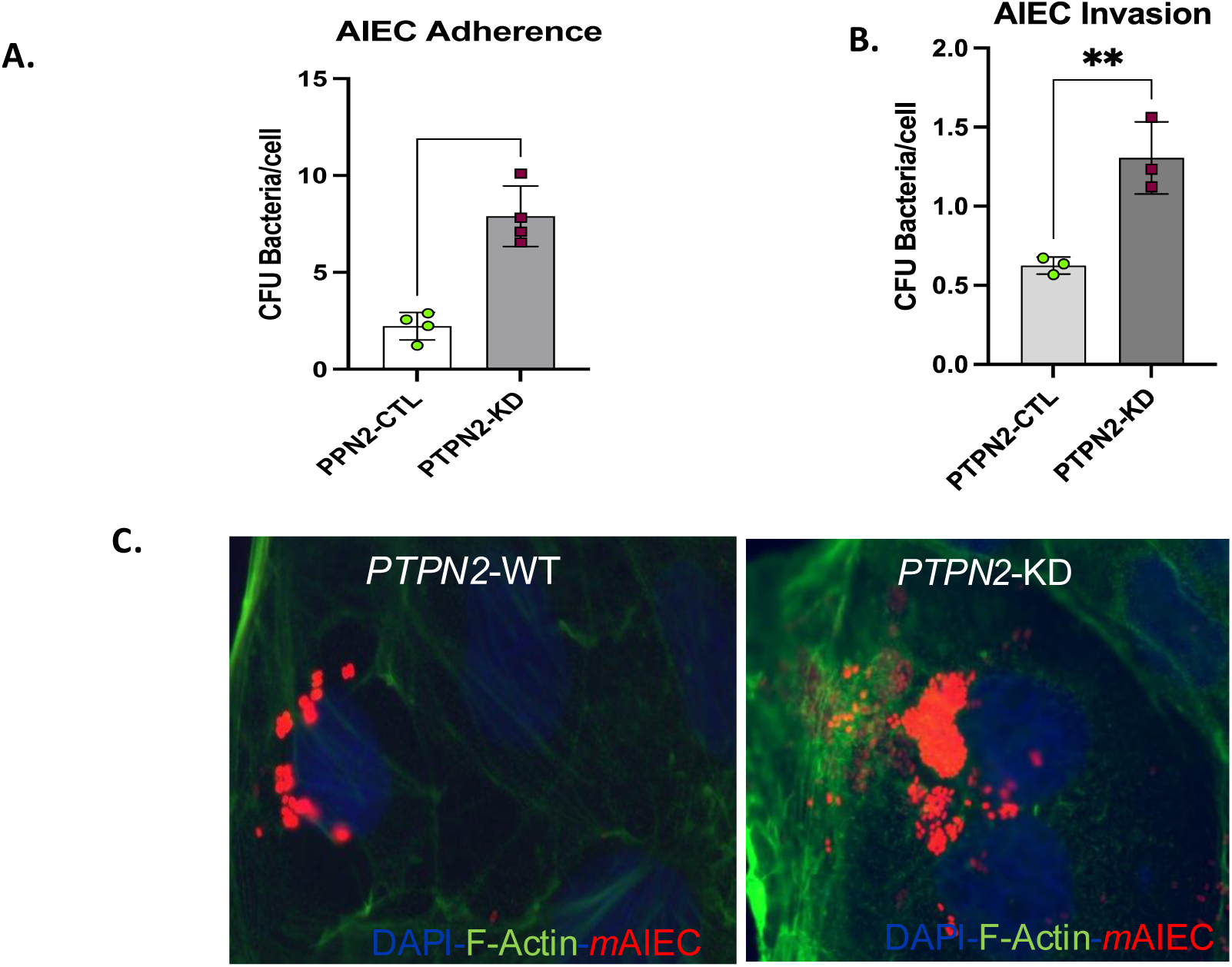
Loss of PTPN2 in IECs increases susceptibility towards AIEC adherence and invasion. Caco-2 BBe cells were transfected with either control shRNA (*PTPN2*-CTL) or PTPN2 targeting shRNA (*PTPN2*-KD). AIEC LF-82 shows higher **A**. Adherence (P<0.001) and **B.** Invasion (P<0.001) of *PTPN2*-KD cells compared to controls. Control and KD Caco-2 BBe cells were seeded on coverslips and infected with *m*AIEC^red^ for 5 hours, washed with PBS and incubated with gentamycin and bacterial invasion was visualized via immunofluorescence microscopy **C.** Immunofluorescence showing increased *m*AIEC^red^ invasion of *PTPN2*-KD cells compared to control.

**Supplementary Figure 2A:**
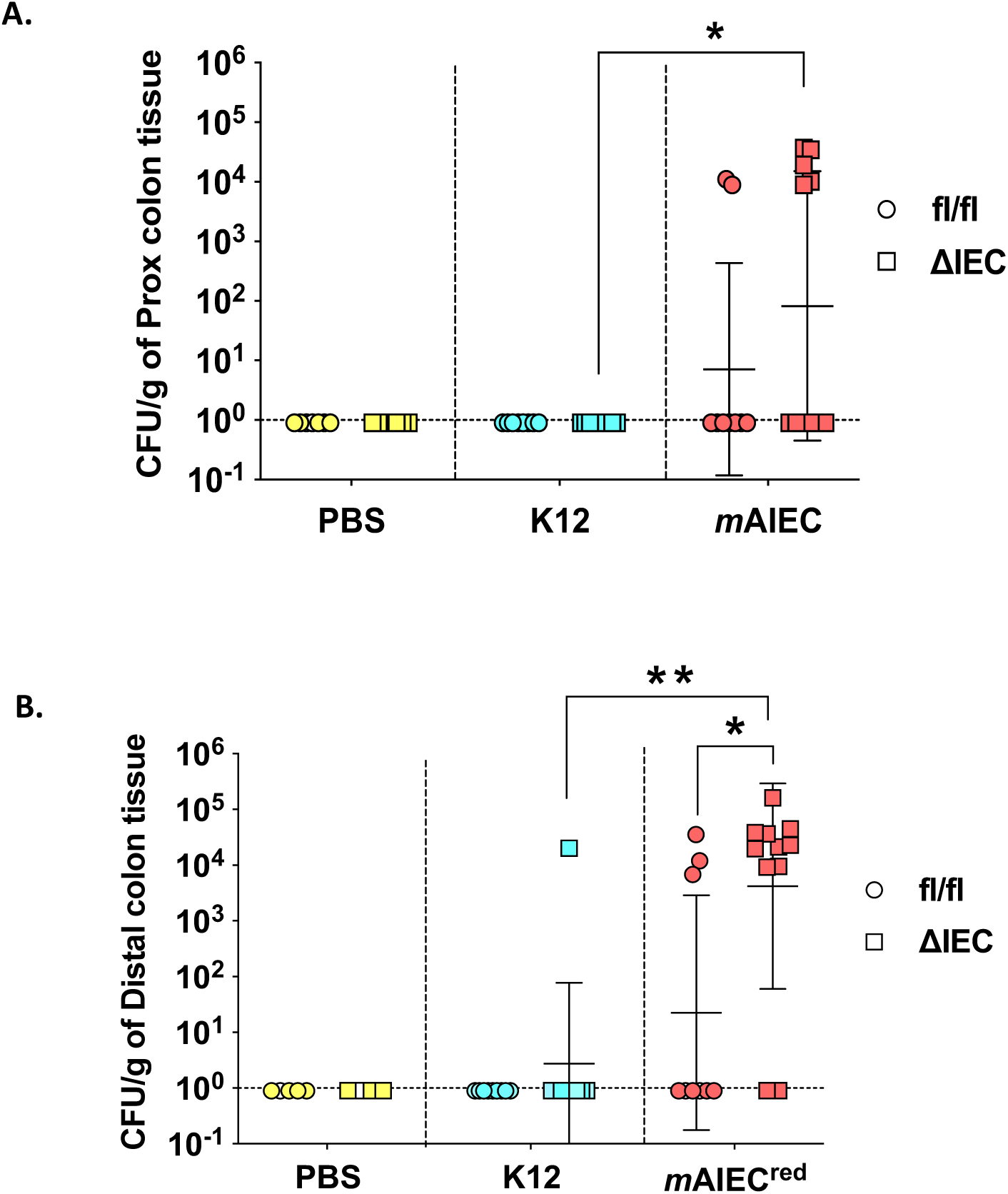
Loss of intestinal epithelial *Ptpn2* promotes *m*AIEC^red^ colonization. (A) Bacterial colonies were enumerated from proximal colon whole tissues, *Ptpn2*^ΔIEC^ - *m*AIEC^red^ mice have greater *m*AIEC^red^ invasion compared to *Ptpn2*^ΔIEC^ – K12 mice (*P*=0.0194). (B) *Ptpn2*^ΔIEC^ mice display higher *m*AIEC^red^ colonization compared to *Ptpn2*^fl/fl^ controls (*P*=0.002).

**Supplementary Figure 3:**
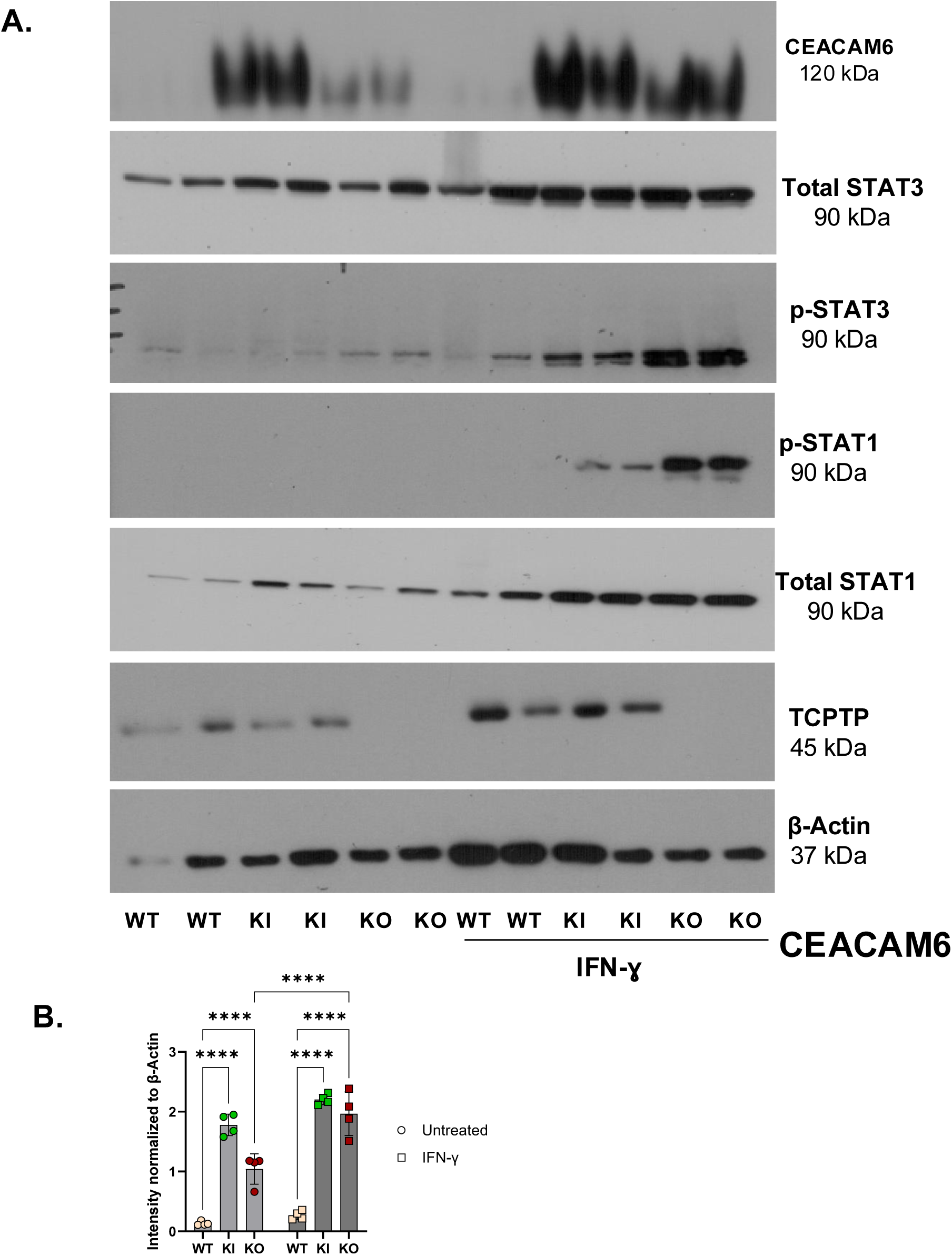
IFN-ɣ Promotes CEACAM6 Expression in *PTPN2*- deficient Caco-2 BBe cells. Caco-2 BBe cells, WT, KI and KO for PTPN2 were treated with IFN-ɣ at 100U/ml for 24 hours. **A.** Representative western blot images depicting β-actin, PTPN2, phosphorylated (p-STAT1/3), STAT(1 and 3) and CEACAM6. p-STAT1 and 3 protein expression in *PTPN2*-KI and PTPN2-KO cells is elevated after IFN-ɣ treatment. CEACAM6 protein expression was elevated at basal states for *PTPN2*-KI and KO cells and was further elevated after IFN-ɣ treatment. **B.** Densitometry analysis of CEACAM6 protein expression after administration of IFN-ɣ. CEACAM6 protein expression is significantly increased in IFN-ɣ treated *PTPN2*-KO cells compared to untreated controls.

**Supplementary Figure 4:**
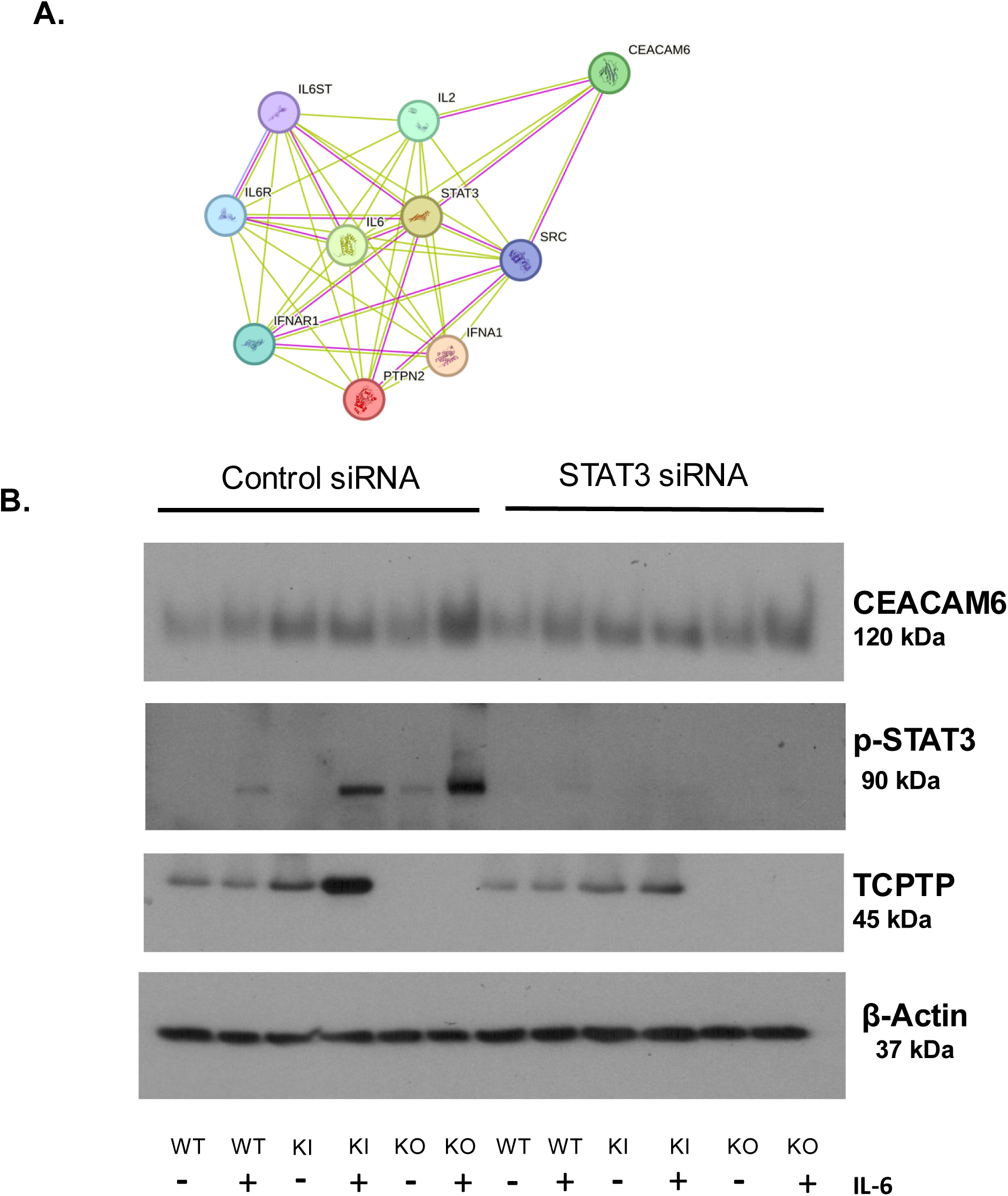
STAT3 silencing does not reduce CEACAM6 Over-expression in PTPN2-KI and KO cell lines. **A.** STRING software analysis shows relation between *PTPN2* protein product-TCPTP, STAT3 and CEACAM6. WT, KI and KO cells were treated with non-targeting control (siControl) and STAT3 specific (siSTAT3) siRNA. After 72 hours, IL-6 was administered. **B.** Representative western blot images showing reduced phosphorylated-STAT3. No appreciable changes in the expression of CEACAM6 was observed between treatment groups or genotypes after silencing of STAT3.

## REFERENCES

1. Jairath, V. and B.G. Feagan, Global burden of inflammatory bowel disease. The Lancet Gastroenterology & Hepatology, 2020. 5(1): p. 2–3.

2. Graham, D.B. and R.J. Xavier, Pathway paradigms revealed from the genetics of inflammatory bowel disease. Nature, 2020. 578(7796): p. 527-539.

3. Mirkov, M.U., B. Verstockt, and I. Cleynen, Genetics of inflammatory bowel disease: beyond NOD2. The lancet Gastroenterology & hepatology, 2017. 2(3): p. 224–234.

4. Strober, W., I.J. Fuss, and R.S. Blumberg, The immunology of mucosal models of inflammation. Annual review of immunology, 2002. 20(1): p. 495–549.

5. Lennard-Jones, J., Classification of inflammatory bowel disease. Scandinavian Journal of Gastroenterology, 1989. 24(sup170): p. 2–6.

6. Lakatos, P.L., Environmental factors affecting inflammatory bowel disease: have we made progress? Digestive Diseases, 2009. 27(3): p. 215–225.

7. Bianco, A.M., M. Girardelli, and A. Tommasini, Genetics of inflammatory bowel disease from multifactorial to monogenic forms. World journal of gastroenterology, 2015. 21(43): p. 12296.

8. De Lange, K.M., et al., Genome-wide association study implicates immune activation of multiple integrin genes in inflammatory bowel disease. Nature genetics, 2017. 49(2): p. 256–261.

9. Barrett, J.C., et al., Genome-wide association study and meta-analysis find that over 40 loci affect risk of type 1 diabetes. Nature genetics, 2009. 41(6): p. 703–707.

10. Scharl, M., et al., Crohn’s disease-associated polymorphism within the PTPN2 gene affects muramyl-dipeptide-induced cytokine secretion and autophagy. Inflammatory bowel diseases, 2012. 18(5): p. 900–912.

11. MR, S., et al., The Clinical Relevance of the IBD-Associated Variation Within the Risk Gene Locus Encoding Protein Tyrosine Phosphatase Non-Receptor Type 2 in Patients of the Swiss IBD Cohort. Digestion, 2016. 93(3).

12. Heinonen, K.M., et al., T-cell protein tyrosine phosphatase deletion results in progressive systemic inflammatory disease. Blood, 2004. 103(9): p. 3457–3464.

13. You-Ten, K.E., et al., Impaired bone marrow microenvironment and immune function in T cell protein tyrosine phosphatase–deficient mice. The Journal of experimental medicine, 1997. 186(5): p. 683–693.

14. Scharl, M., et al., Protection of epithelial barrier function by the Crohn’s disease associated gene, protein tyrosine phosphatase N2. Gastroenterology, 2009. 137(6): p. 2030–2040 e5.

15. Krishnan, M. and D.F. McCole, T cell protein tyrosine phosphatase prevents STAT1 induction of claudin-2 expression in intestinal epithelial cells. Ann N Y Acad Sci, 2017. 1405(1): p. 116–30.

16. Simoncic, P.D., et al., The T cell protein tyrosine phosphatase is a negative regulator of janus family kinases 1 and 3. Current biology, 2002. 12(6): p. 446–453.

17. Hodge, J.A., et al., The mechanism of action of tofacitinib-an oral Janus kinase inhibitor for the treatment of rheumatoid arthritis. Clin Exp Rheumatol, 2016. 34(2): p. 318–328.

18. Panés, J., et al., Tofacitinib for induction and maintenance therapy of Crohn’s disease: results of two phase IIb randomised placebo-controlled trials. Gut, 2017. 66(6): p. 1049–1059.

19. Sandborn, W.J., et al., Tofacitinib as induction and maintenance therapy for ulcerative colitis. New England Journal of Medicine, 2017. 376(18): p. 1723–1736.

20. Jostins, L., et al., Host–microbe interactions have shaped the genetic architecture of inflammatory bowel disease. Nature, 2012. 491(7422): p. 119–124.

21. Tamboli, C.P., et al., Dysbiosis in inflammatory bowel disease. Gut, 2004. 53(1): p. 1–4.

22. Martinez-Medina, M., et al., Molecular diversity of Escherichia coli in the human gut: new ecological evidence supporting the role of adherent-invasive E. coli (AIEC) in Crohn’s disease. Inflammatory bowel diseases, 2009. 15(6): p. 872–882.

23. Barnich, N., et al., Regulatory and functional co-operation of flagella and type 1 pili in adhesive and invasive abilities of AIEC strain LF82 isolated from a patient with Crohn’s disease. Molecular microbiology, 2003. 48(3): p. 781–794.

24. Eaves-Pyles, T., et al., Escherichia coli isolated from a Crohn’s disease patient adheres, invades, and induces inflammatory responses in polarized intestinal epithelial cells. International Journal of Medical Microbiology, 2008. 298(5-6): p. 397–409.

25. Desilets, M., et al., Genome-based definition of an inflammatory bowel disease-associated adherent-invasive Escherichia coli pathovar. Inflammatory bowel diseases, 2016. 22(1): p. 1–12.

26. Wen, W., et al., PUFAs add fuel to Crohn’s disease-associated AIEC-induced enteritis by exacerbating intestinal epithelial lipid peroxidation. Gut Microbes, 2023. 15(2): p. 2265578.

27. Xu, Y., et al., Crohn’s disease-associated AIEC inhibiting intestinal epithelial cell-derived exosomal let-7b expression regulates macrophage polarization to exacerbate intestinal fibrosis. Gut Microbes, 2023. 15(1): p. 2193115.

28. Shaler, C.R., et al., Psychological stress impairs IL22-driven protective gut mucosal immunity against colonising pathobionts. Nature Communications, 2021. 12(1): p. 6664.

29. Elhenawy, W., C.N. Tsai, and B.K. Coombes, Host-specific adaptive diversification of Crohn’s disease-associated adherent-invasive Escherichia coli. Cell Host & Microbe, 2019. 25(2): p. 301–312. e5.

30. Horowitz, A., et al., Paracellular permeability and tight junction regulation in gut health and disease. Nature Reviews Gastroenterology & Hepatology, 2023: p. 1–16.

31. Turner, J.R., Intestinal mucosal barrier function in health and disease. Nature reviews immunology, 2009. 9(11): p. 799–809.

32. Barnich, N., et al., CEACAM6 acts as a receptor for adherent-invasive E. coli, supporting ileal mucosa colonization in Crohn disease. The Journal of clinical investigation, 2007. 117(6): p. 1566–1574.

33. Shawki, A., et al., The autoimmune susceptibility gene, PTPN2, restricts expansion of a novel mouse adherent-invasive E. coli. Gut Microbes, v. 11(6): p. 1547–1566.

34. Marchelletta, R.R., et al., T cell protein tyrosine phosphatase protects intestinal barrier function by restricting epithelial tight junction remodeling. The Journal of clinical investigation, 2021. 131(17).

35. Knights, D., et al., Complex host genetics influence the microbiome in inflammatory bowel disease. Genome Med 6: *107*. 2014.

36. Yilmaz, B., et al., The presence of genetic risk variants within PTPN2 and PTPN22 is associated with intestinal microbiota alterations in Swiss IBD cohort patients. PloS one, 2018. 13(7): p. e0199664.

37. Holmer, R., et al., Interleukin-6 trans-signaling increases the expression of carcinoembryonic antigen-related cell adhesion molecules 5 and 6 in colorectal cancer cells. BMC cancer, 2015. 15: p. 1–12.

38. Camprubí-Font, C. and M. Martinez-Medina, Why the discovery of adherent-invasive Escherichia coli molecular markers is so challenging? World journal of biological chemistry, 2020. 11(1): p. 1.

39. Abdelhalim, K.A., A. Uzel, and N.G. Ünal, Virulence determinants and genetic diversity of adherent-invasive Escherichia coli (AIEC) strains isolated from patients with Crohn’s disease. Microbial pathogenesis, 2020. 145: p. 104233.

40. Spalinger, M.R., et al., Autoimmune susceptibility gene PTPN2 is required for clearance of adherent-invasive Escherichia coli by integrating bacterial uptake and lysosomal defence. Gut, 2022. 71(1): p. 89–99.

41. Douadi, C., et al., Anti-TNF agents restrict Adherent-invasive Escherichia coli replication within macrophages through modulation of Chitinase 3-like 1 in patients with Crohn’s disease. Journal of Crohn’s and Colitis, 2022. 16(7): p. 1140–1150

